# Chronic pain gene expression changes in the brain and relationships with clinical traits

**DOI:** 10.1101/2022.12.28.22283994

**Authors:** Keira JA Johnston, Alanna C. Cote, Emily Hicks, Jessica Johnson, Laura M. Huckins

## Abstract

**Background:** Chronic pain is a common, poorly-understood condition. Genetic studies including genome wide association studies (GWAS) identify many relevant variants, which have yet to be translated into full understanding of chronic pain. Transcriptome wide association study using transcriptomic imputation (TI) methods such as S-PrediXcan can help bridge this genotype-phenotype gap.

**Methods:** We carried out TI using S-PrediXcan to identify genetically regulated gene expression (GREX) in thirteen brain tissues and whole blood associated with Multisite Chronic Pain (MCP). We then imputed GREX for over 31,000 Mount Sinai Bio*Me*™ participants and performed phenome-wide association study (PheWAS) to investigate clinical relationships in chronic pain associated gene expression changes.

**Results:** We identified 95 experiment-wide significant gene-tissue associations (p<7.97×10^−7^), including 35 unique genes, and an additional 134 gene-tissue associations reaching within-tissue significance, including 53 additional unique genes. Of 89 unique genes total, 59 were novel for MCP and 18 are established drug targets. Chronic pain GREX for 10 unique genes was significantly associated with cardiac dysrhythmia, metabolic syndrome, disc disorders/ dorsopathies, joint/ligament sprain, anemias, and neurological disorder phecodes. PheWAS analyses adjusting for mean painscore showed associations were not driven by mean painscore.

**Conclusions:** We carried out the largest TWAS of any chronic pain trait to date. Results highlight potential causal genes in chronic pain development, and tissue and direction of effect. Several gene results were also drug targets. PheWAS results showed significant association for phecodes including cardiac dysrhythmia and metabolic syndrome, indicating potential shared mechanisms.

## Introduction

Chronic pain is a common, debilitating condition (1–3). Risk factors and mechanisms of chronic pain development are not fully understood. Treating chronic pain successfully is a complex process; many pharmacological treatments are suboptimal and associated with serious risks such as addiction and dependence (4), as seen in the opioid epidemic in the United States (5–7), with similar trends seen in the United Kingdom (8, 9).

Genetic studies into chronic pain (10–12) and conditions associated with significant chronic pain (e.g. rheumatoid arthritis (13), endometriosis (14), and migraine (15)) have revealed hundreds of genetic loci of interest, but these results have not been translated into actionable treatment for chronic pain. In the pathway from genotype to phenotype, transcription and gene expression represent intermediate steps. Understanding expression changes associated with chronic pain could aid in further understanding of the mechanisms and best pharmaceutical treatments for chronic pain.

Transcriptomic Imputation (TI) approaches combine expression quantitative trait loci (eQTL) and genome wide association study (GWAS) association statistics to identify trait-associated genetically regulated gene expression (GREX), providing directional and tissue-specific context (16–19). This approach is especially useful since changes to the brain and spinal cord have been widely implicated in the development of chronic pain (20–23), and brain tissue is relatively inaccessible and impossible to assay in living study participants. TI studies have been carried out in a range of conditions (24–26), including complex traits that are commonly associated with chronic pain (e.g., rheumatoid arthritis (27), Crohn’s disease (28, 29), lupus (30), and migraine (15)) but no direct TI analyses of chronic pain have been undertaken. In this study, we apply a TI method, S-PrediXcan (17), to impute GREX in thirteen brain regions, and test for association with multisite chronic pain (10).

As outlined above, GWAS have revealed hundreds of SNP associations, but there is an outstanding need to interrogate the consequences of these genetic variants in clinical data (31, 32). Phenome-wide association studies (PheWAS) represent a way to test for significant associations between exposures (which can be genetic variants, biomarkers, or other risk factors) and large sets of phenotypes, such as anthropomorphic trait values, blood lipid levels, or ICD-10 or other electronic-health record traits (33). Previous PheWAS analyses have created disease-disease networks and shown relationships between chronic pain conditions, including the relationship between seronegative rheumatoid arthritis and fibromyalgia (34), and between genetic risk for problematic opioid use and pain-related phenotypes (35). Here, we tested for association between chronic pain-associated GREX and a phenome composed of over 1,000 phecodes in an ancestrally diverse hospital biobank.

This study involves GWAS summary statistics from one of the largest studies chronic pain to date, where chronic pain was examined as a complex disease trait (10). This may represent a more powerful and tractable way to uncover genetic variation specific to chronic pain development in comparison to genetic study of chronic pain associated conditions. We highlight genes of interest through their genetically regulated gene expression, in specific tissues, relevant to mechanisms of chronic pain development. We also present the first PheWAS of genetically-regulated gene expression for chronic pain.

## Methods and Materials

### GWAS output and phenotype: multisite chronic pain

GWAS output from a 2019 study of multisite chronic pain (MCP) (10), a chronic pain phenotype quantifying the number of sites of chronic pain on the body, was used in this study. MCP was found to be a complex, polygenic chronic pain trait, associated with validated chronic pain trait Chronic Pain Grade, and genetically correlated with psychiatric and other disorders. Studying chronic pain as a disease trait, as opposed to studying conditions associated with chronic pain, is in line with recent changes to ICD-11 coding for chronic pain and International Association for the Study of Pain (IASP) definitions of chronic pain (36–38). Considering chronic pain in this way may provide greater insight into pain-associated genetic variation. Findings are described in full in the original publication, but genes involved in development and maintenance of the CNS, and immune processes, were found to be associated with multisite chronic pain using MAGMA (a gene-level association testing method) (39), and gene expression of MCP-related genes was enriched in the brain, particularly in frontal cortex. To further understand mechanisms of chronic pain development, GWAS summary statistics were used to perform TWAS analysis via a transcriptomic imputation approach using S-PrediXcan (17).

### Discovery of genetically-regulated gene expression in chronic pain

Genetically regulated gene expression (GREX) was imputed using MCP GWAS output and TI models from the Genotype-Tissue Expression Project (GTEx) (40) in 13 brain tissues (amygdala, anterior cingulate cortex BA24, caudate basal ganglia, cerebellar hemisphere, cerebellum, cortex, frontal cortex BA9, hippocampus, hypothalamus, nucleus accumbens basal ganglia, putamen basal ganglia, spinal cord cervical c-1, and substantia nigra) and whole blood, using S-PrediXcan. Multiple testing correction was performed using Bonferroni-correction, resulting in two thresholds for significance: (1) a per-tissue threshold correcting for all genes tested in each tissue (Table 1) (2) an experiment-wide threshold, correcting for all genes tested across all tissues (p=7.9e-7).

### Downstream analysis: FUMA

Pathway analyses were carried out using FUMA GENE2FUNC (41) including all per-tissue significant gene results (N = 89). We tested for enrichment of all gene sets available in FUMA GENE2FUNC with all genes that had at least one S-PrediXcan prediction model available and were included in FUMA as ‘background’ (N = 15, 588 background genes compared to FUMA default N = 20260 protein-coding background genes). Genes listed as drug targets and found to be significant in S-PrediXcan were investigated using FUMA DrugBank lookup results.

### Phenome-wide association analysis in Mount Sinai Bio*Me™*

In order to probe relationships between our MCP-associated GREX and clinical phenotypes, we performed a series of phenome-wide association studies (PheWAS) in the Mount Sinai Bio*Me™* biobank.

Bio*Me™* is a large, diverse, hospital-based biobank, including electronic health record (EHR) and genotype data for 31, 704 participants in the first data freeze. A total of 1, 236 phecodes for Bio*Me™* participants were included in analyses in this paper. Phecodes are a high-throughput phenotyping method for electronic health records, where ICD-10 codes are manually grouped according to clinical similarity, reducing EHR dimension and complexity (42). Here, we use previously curated phecodes (43).

First, we imputed MCP-GREX (chronic pain-related genetically regulated gene expression) for 31, 704 Bio*Me™* freeze 1 participants, split across six genotype-derived ancestry groups (Table 2). Specifically, we imputed GREX in all 13 brain regions and in whole blood for all 89 unique genes previously identified as significant MCP-GREX. We tested for association between these GREX values and Bio*Me™* phecodes with at least ten available cases in at least one ancestry (total N phecodes = 1,236 (43)). Results were meta-analyzed using inverse variance weighted meta-analysis in METAL (44). Multiple testing correction was then applied using a within-gene false discovery rate (FDR) procedure.

To validate our MCP-associations, we tested whether MCP-associated genes were associated with pain in this cohort. A numeric rating scale (NRS) 0-10, where 10 is the worst pain possible and 0 is no pain, was recorded for Bio*Me™* participants and was aggregated into a mean pain score. Associations were tested between significant MCP-GREX results and mean pain score across the six ancestry groups, results were meta-analysed, and FDR multiple testing correction within gene was performed.

## Results

### Novel brain-specific genes and pathways associated with chronic pain identified with transcriptomic imputation

We applied S-PrediXcan to the largest available summary statistics for multi-site chronic pain (N=387,649). We identified 95 experiment-wide significant gene-tissue associations (p<7.97×10^−7^), including 35 unique genes associated across 14 brain regions (Table 3). Since an experiment-wide threshold is likely overly conservative given the degree of shared eQTLs between tissues, we also applied a within-tissue Bonferroni threshold (Table 1, Figure 1A & 1B). We identified an additional 134 gene-tissue associations reaching within-tissue significance, including an additional 53 additional unique genes associated with multi-site chronic pain across 14 brain regions.

**Figure 1:**
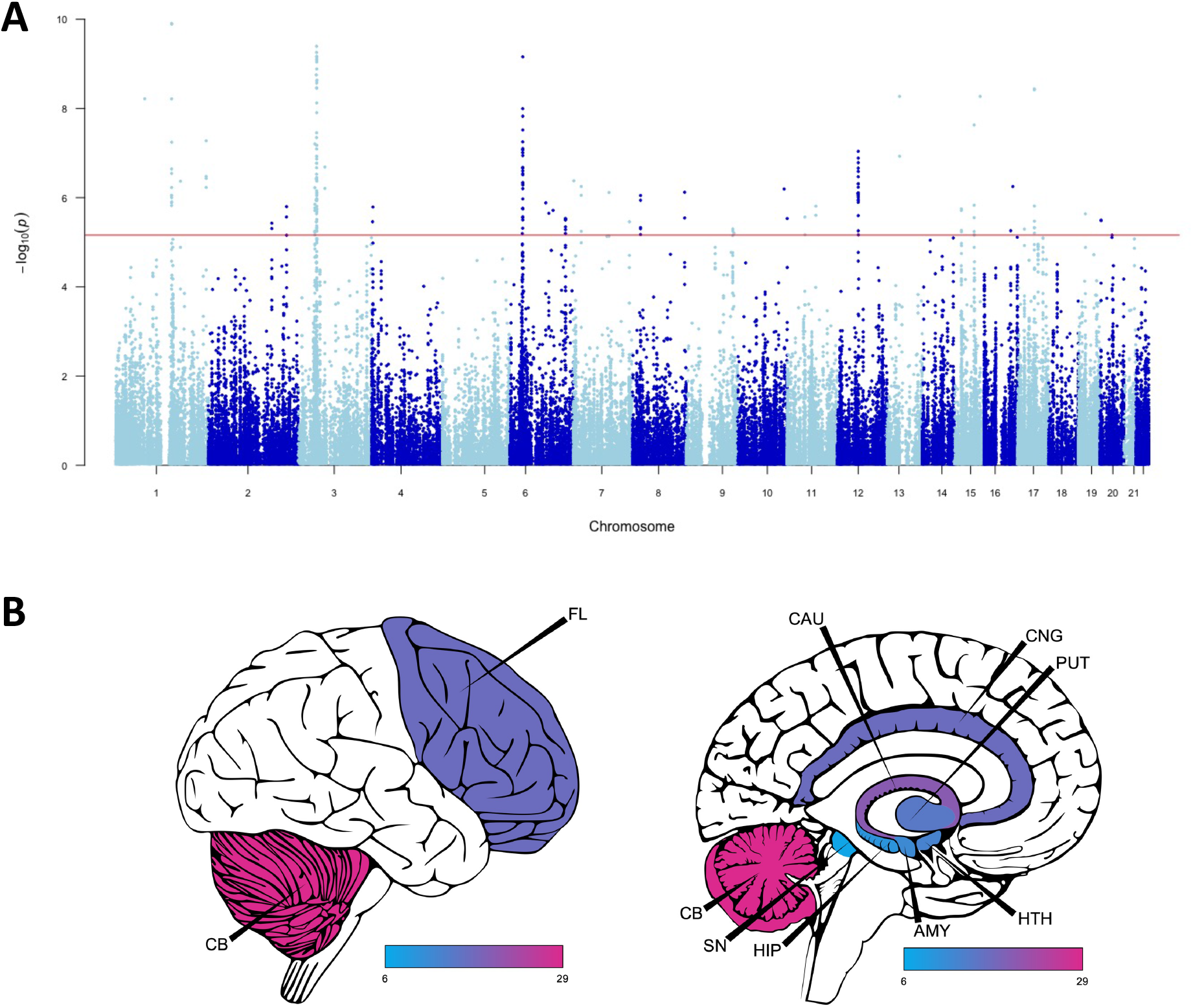
S-PrediXcan analysis identifies 89 unique genes associated with chronic pain. A: S-Predixcan analyses identified 89 unique significant gene associations across fourteen tissues. Red line indicates most conservative per-tissue significance threshold. B: Number of significant MCP-GREX genes per brain region. CB = cerebellum, FL = frontal lobe, SN = substantia nigra, HIP = hippocampus, AMY = amygdala, HTH = hypothalamus, PUT = putamen, CNG = anterior cingulate cortex, CAU = caudate. Created using Cerebroviz (45).

Of these 89 genes, 59 were not previously associated with multisite chronic pain (10). Of these genes, 49 were associated only in a single tissue - for example MCP-GREX of CDK14 (cyclin-dependent kinase 14) is found only in the cortex. MCP-GREX of CEP170, MON1B, MST1, RP11-160H22.5, URM1, VPS33B, ZNF197, and ZNF35 was found only in whole blood. In contrast, several genes show MCP-GREX across a wide range or in some cases across all tested tissues – GMPPB MCP-GREX was found in all brain tissues and whole blood, MCP GREX of RBM6 in 13 of 14 tissues, RPS26 MCP-GREX in 12 tissues, UHRF1BP1 in 11 tissues, and SUOX in 10 tissues.

In order to test whether significant associations were enriched in any specific brain region, we performed binomial tests for enrichment, comparing the proportion of experiment-wide significant associations identified in a brain region to the proportion of genes tested in that brain region. We found significantly more experiment-wide significant associations in the nucleus accumbens basal ganglia than might be expected by chance (14.7% vs. 7.6%, p_Binomial_=0.0075), and significantly fewer than might be expected in the cerebellar hemisphere (4.2% vs. 9.0%; p_Binomial_=0.038). Repeating this test for nominally associated genes identified three brain regions with fewer associations than expected by chance; hippocampus (5.3% vs. 5.8%, p_Binomial_=0.033), spinal cord cervical c1 (4.4% vs. 5.1%, p_Binomial_=0.0014), substantia nigra (3.4% vs. 4.0%, p_Binomial_=0.0035).

### Downstream analyses indicate potential chronic pain drug targets

To identify functional patterns of MCP-GREX associations, we conducted a gene-set enrichment analysis using FUMA. Genes associated with MCP-GREX were significantly enriched in the positional gene set chr3p21 (p = 5.27 ×10^−19^; Figure 2a), a locus previously implicated in anorexia nervosa (46). In addition, MCP-GREx genes were significantly enriched for genes associated with eight GWAS (Figure 2b). As a positive confirmation, this included a previous GWAS of MCP ((10), p=5.54×10^−6^). The other seven enriched GWAS included sleep duration (short sleep) (p=2.27×10^−11^), extremely high intelligence (p=6.66 ×10^−8^), regular attendance at gyms and sports clubs (p=6.66×10^−8^) and religious groups (p=7.66 ×10^−6^), as well as inflammatory conditions (ulcerative colitis, p=1.95 ×10^−5^, IBD, p=5.9 ×10^−3^), and age at first birth (p=1.57 ×10^−3^). FUMA DrugBank lookups (Table 4) identified 19 genes that are known drug targets, with a wide range of drug categories implicated including nutritional supplements, monoclonal antibodies, tyrosine kinase inhibitors, and anti-cancer compounds.

**Figure 2:**
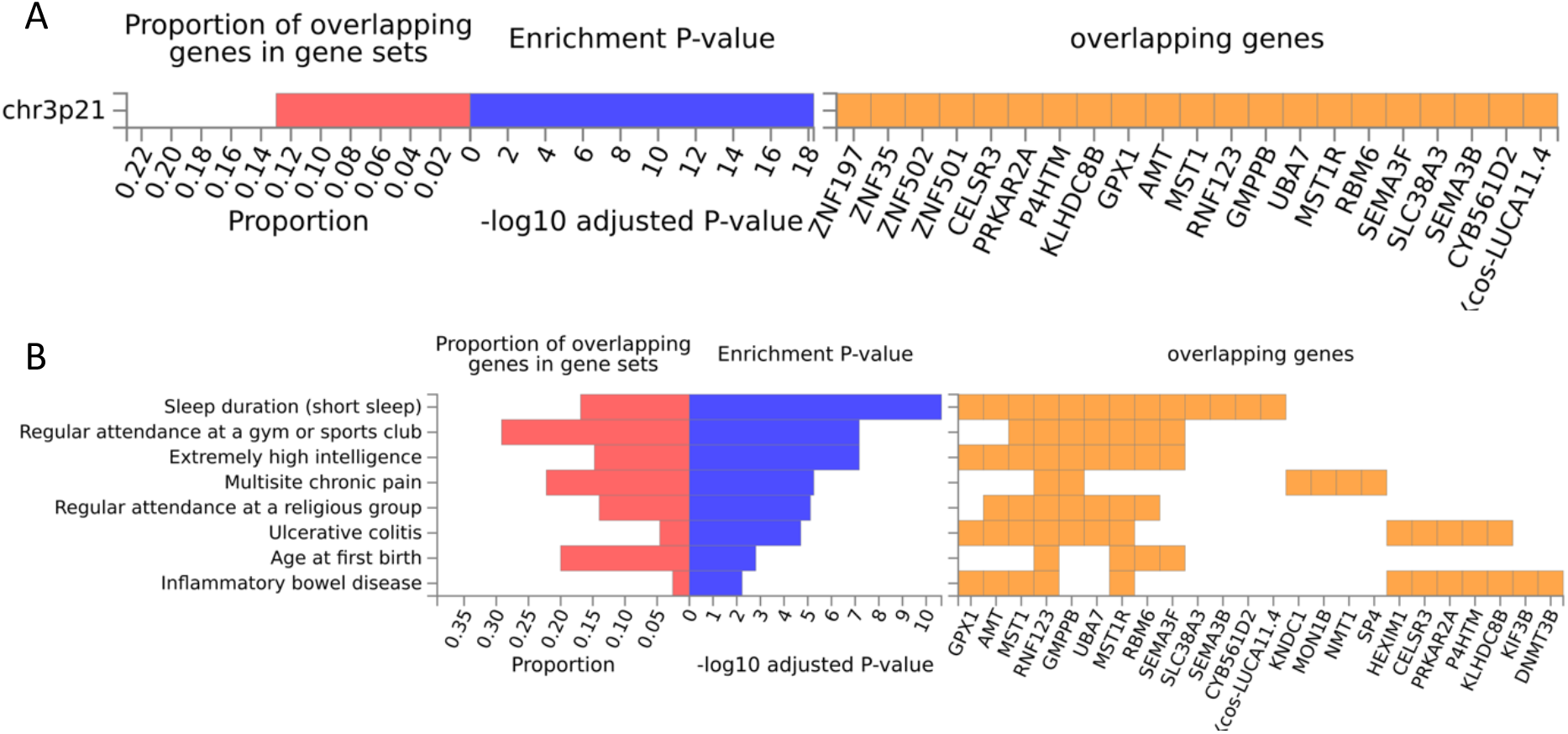
Gene set enrichment analysis identifies positional and GWAS enrichments. A: FUMA gene set enrichment identified one positional gene set (chr3p21) enriched for MCP-GREX genes. B: Enrichment analyses showed nine GWAS catalog traits significantly enriched for MCP-GREX genes.

### Clinical associations with chronic pain genetically regulated gene expression revealed through PheWAS

To probe potential clinical consequences of our MCP-associated genes, we performed a phenome-wide association study (PheWAS) in the Mount Sinai Bio*Me™* biobank. First, we imputed MCP-GREX for 89 significant MCP-GREX gene-tissue associations for 18,806 biobank participants who had available mean pain score data, and tested for association between GREX and mean pain score. We identified 37 associations including 10 unique genes between MCP-GREX and mean pain score including 10/89 MCP-GREX genes (Table 5). Next, we tested for phenome-wide associations. We identified 16 GREx-phecode associations across nine brain regions, including ten unique gene-phecode associations (Table 6; Figure 3). Associated phecodes included cardiac dysrhythmia, metabolic syndrome, disc disorders/ dorsopathies, joint/ligament sprain, anemias, and neurological disorders.

**Figure 3:**
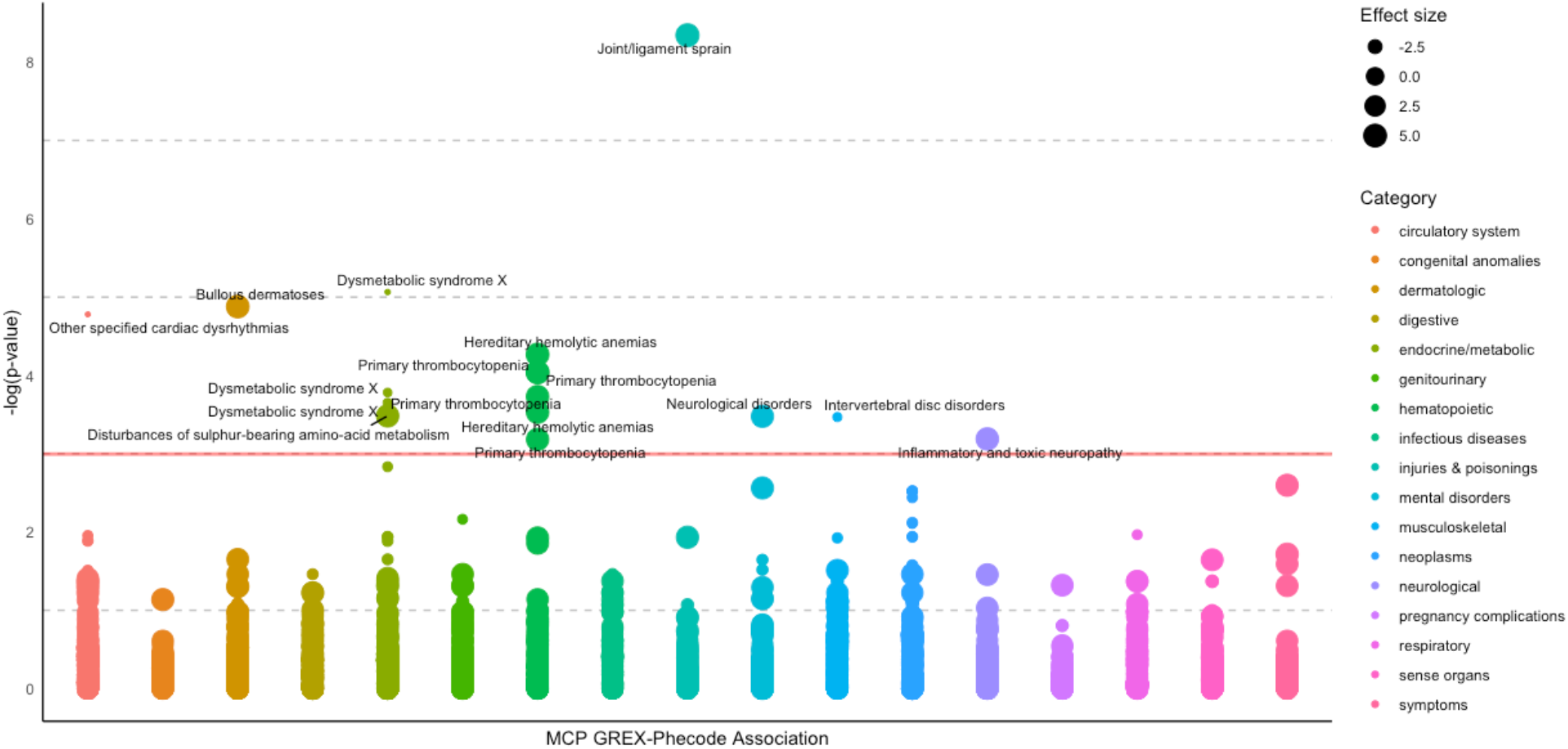
Phenome-wide associations with chronic pain associated genes. Effect size = z score value for association between MCP-GREX and phecode. Red horizontal line indicates significance threshold (-log10(0.05) = 2.995), phecodes are colour coded according to wider phecode category (using mapping tables made available at https://phewascatalog.org/phecodes_icd10 and associated with Wu & Denny 2019).

As pain and chronic pain are core symptoms of many of these diagnoses, and some genes with significant MCP-GREX were significantly associated with pain NRS, it is difficult to discern whether our MCP genes are associated with pain, or directly with the trait itself. Therefore, we repeated our PheWAS, including mean pain scores derived from pain NRS information as covariates. We also carried out a PheWAS with identical adjustment to our main analyses (no adjustment for mean pain score) on the same subset of individuals included in pain score-adjusted PheWAS analyses. We found results to be significantly different from main PheWAS results, but after comparison to the unadjusted-subset PheWAS, this appears to be driven by a reduction in sample size rather than by mean pain score (Supplementary Tables 1 & 2). Sample size is significantly reduced in analyses with adjustment for pain score as many Bio*Me™* participants do not have pain NRS information available.

## Discussion

These results reveal novel genes relevant to chronic pain development, providing new insight into mechanisms of chronic pain, and a set of genes theoretically enriched for causal effect in chronic pain. Through applying TI using S-PrediXcan to multisite chronic pain GWAS output, we can perform a well-powered study of gene expression in brain tissue and whole blood, the likes of which are currently not feasible with existing cohorts where chronic pain phenotyping, genotype and expression data are available together due to limited sample sizes.

### Gene findings give insight on shared pathways between chronic pain other medical conditions

GREX of ILRUN, a gene involved in innate immune response and highly expressed in B cells (47), is significantly associated with MCP in basal ganglia of the nucleus accumbens, hypothalamus, amygdala, and cortex in the original S-PrediXcan analysis, and with primary thrombocytopenia across all four tissues in our PheWAS. Primary thrombocytopenia is an autoimmune disorder where platelets are destroyed by self-reacting antibodies, resulting in low peripheral plate counts and symptoms such as joint and abdominal pain, bleeding, and bruising. In addition, ILRUN has been linked to the renin-angiotensis-aldosterone system (RAAS) (involved in blood volume, sodium reabsorption, and vascular tone amongst other processes) in a study on SARS-COV2 infection (48). The RAAS has also been implicated in chronic pain, as peripheral small Aδ and C fibres which transmit pain signals contain cells expressing RAAS components, and RAAS modulators have been shown to have an effect on pain relief (49) – results here suggest a role for ILRUN in the brain in chronic pain development, in addition to this relationship with pain perception in the periphery.

MCP-GREX of MON1B in both amygdala and cervical spinal cord c-1 was found to be significantly associated with anemias – this phecode encompasses conditions including sickle-cell anemia and thalassemia, in addition to haemolytic anemias, chronic conditions which are often associated with significant pain (50). Iron deficiency and iron-deficiency anemia are also generally associated with chronic inflammatory disease, which again in turn is often associated with chronic pain (51). In addition, dysregulation of iron metabolism in general can play a key role in homeostasis of immune cells, and inflammatory processes (52, 53). MON1B encodes a protein involved in phagosome maturation and removal of apoptotic cell remains, with defects associated with autoimmune pathology (54), which has been shown to play a significant role in chronic pain (55). The protein encoded by MON1B is also a key part of the regulation of endocytic sorting by Numb, and so is linked to cell migration, asymmetric cell division, and differentiation (56).

DCAKD encodes a protein that may be involved in neurodevelopment (57) and is expressed widely in the brain (58) – MCP-GREX of this gene in the caudate basal ganglia was negatively associated with cardiac dysrhythmia in PheWAS analyses. Previous studies indicate a relationship between some MRI markers of cerebral small vessel disease and this gene (59) and Friedrich’s ataxia (60), a disease of progressive neurodegeneration, movement disorder, and heart and spinal problems (61, 62). Respiratory sinus arrhythmia (heart rate variability), is thought to represent (potentially inappropriate) hyperarousal and has been linked to emotion regulation and chronic pain (63, 64). In addition, certain nerve blocks have applications in treating both cardiac and chronic pain conditions (65).

ECM1 encodes a protein involved in type 2 helper T cell migration (66), and potentially involved in skin development (67). In PheWAS analyses ECM1 MCP-GREX in three different brain tissues was associated with dysmetabolic syndrome X (aka metabolic syndrome) – metabolic syndrome is defined as 3 or more of: elevated waist circumference, elevated triglycerides, reduced HDL cholesterol, elevated BP and elevated fasting plasma glucose (68, 69). This syndrome has been associated with increased risk of cardiovascular disease and type 2 diabetes (68, 70). T cells have been associated with development of insulin resistance in obesity (71), and having metabolic syndrome can affect T cell development (reviewed by (72)), and amount of memory T cells has been associated with a generally more pro-inflammatory state (73). These cell types have also been explored as factors in chronic pain development and therapeutic targets in chronic pain treatment (74–77). T cells may also represent a sex-dimorphic method by which pain hypersensitivity is mediated (reviewed by (78, 79)).

PACSIN3 encodes a protein involved in the actin cytoskeleton and formation of vesicles (80). This protein also binds TRPV4, an ion channel protein – channelopathy mutations in the TRPV4 gene lead to skeletal dysplasias, charcot-marie tooth disease subtype 2C, premature osteoarthritis, and neurological disorders (81). TRPV4 channels are also important in skin function(82), and are involved in the itch-scratch cycle (83, 84). TRP channels have also been implicated in chronic low back pain (85), and investigated as a therapeutic target in conditions such as fibromyalgia (86, 87). PACSIN3 MCP-GREX in the basal ganglia of the nucleus accumbens was significantly associated with bullous dermatoses in PheWAS analyses. Bullous dermatoses are autoimmune skin conditions where the main symptom is painful blistering (88–91). Although itch and pain are considered overall to be distinct processes (84), they share many similarities (92), and results here suggest the ion channel encoded by TRPV4, and its interaction with the protein encoded by PACSIN3, could be a point of overlap in chronic pain and itch.

RAD51 encodes a protein involved in DNA repair (93, 94), and mutations in this gene have been linked to congenital mirror movement disorder (95) and cancers (96). MCP-GREX at this gene in substantia nigra was significantly associated with disturbances of sulphur bearing amino acid metabolism. This phecode includes conditions such as homocystinuria, where the body is unable to process the amino acid methionine, and Methylenetetrahydrofolate Reductase (MTHFR) Deficiency, where homocysteine levels become elevate (97). These processes are both part of DNA metabolism (98).

Elevated homocysteine levels have been associated with a range of illness and these levels can have toxic effects on the nervous system (99) and negative impact on DNA repair. RAD51 foci (an indicator a cell is under replication stress) (100) were found to be increased in experiments examining folate deficiency (101). Previous studies of elevated homocysteine in rodents also found a causal relationship between this metabolic dysfunction and mechanical allodynia (102) – PheWAS results indicate a potential role for this mechanism of sensitization in human chronic pain too.

SCAI (Suppressor of Cancer Cell Invasion) encodes a transcriptional co-factor involved in regulating invasive cell migration (103), including in gliomas (a tumor type usually found in the brain or spinal cord) (104). MCP-GreX of this gene in the cortex was associated with toxic/ inflammatory neuropathy in PheWAS analyses, and this gene has been found to be differentially expressed in rat spinal cord in models of diabetic neuropathy (105). Our findings indicate a possible similar role for human SCAI in neuropathy. SLC38A3 encodes a glutamine transporter (106), involved in ammonia detoxification and cell energy metabolism amongst other processes. Glutamine is the preferred energy source for rapidly proliferating cell populations such as those in the nervous system, immune system, and cancer cells (107–111). In addition to expression in the brain, SLC38A3 is also expressed in muscle - significant MCP-GREX in caudate basal ganglia was found to be associated with joint and muscle sprain, suggesting the glutamine transporter encoded by SLC38A3 has a central as well as peripheral role during injury. SLC38A3 MCP-GREX in the same brain area was also significantly associated with neurological disorders, in line with research showing relationships between glutamine metabolism in the brain and neurological conditions including traumatic brain injury, dementia, Parkinson disease, and epilepsy (112–115). Gabaergic gene regulatory elements have also been implicated in neurological and psychiatric diseases (116–120), glutamate receptors in neurological dysfunction (121), and treating neurodegeneration through targeting glutamate transporters (122). Activity-dependent synaptic plasticity also involves glutamate and glutamine metabolism (123). Glutamine and other neurometabolites in the glutamine metabolism pathway have also been investigated as potential biomarkers of chronic pain, as concentrations of these factors vary in those with chronic pain compared to controls (124, 125), and glutamine supplementation may be helpful in the treatment of vaso-occlusive crisis in sickle cell disease (126). Glutamine levels have also been associated with individual pain sensitivity differences (127), and migraine (128). Finally, glutamine supplementation has been shown in a small study to reduce opioid use in those with sickle cell, highlighting its potential as a harm (as well as pain) reducing compound in chronic pain treatment (129).

Finally, ERICH2 MCP-GREX in the amygdala was significantly associated with dorsopathies (conditions where there is interruption of/ deviation from normal structure or normal function of the spine). This brain region has been previously associated with chronic pain (130–133), particularly the aspect of pain-related fear (134, 135).

### Comparison with genetic correlation results

Phecodes significantly associated with MCP-GREX included metabolic syndrome, joint and ligament sprains and dorsopathies, and neurological disorders – however, psychiatric disorder related phecodes, and phecodes assigned to chronic pain conditions e.g., rheumatoid arthritis or endometriosis, were not significantly associated with MCP-GREX. This is in contrast to genetic correlation results between MCP and e.g. major depressive disorder, and MCP and rheumatoid arthritis found in a previous study (10). The fact that genetic correlations are calculated using all SNP associations genome-wide rather than at a gene-level, and include genetic variants associated with very small effect in trait variation, may explain these differences in PheWAS results and previous genetic correlation analyses. In addition, S-PrediXcan results in theory represent gene expression changes that occur before chronic pain development (whereas GWAS summary statistics used in LD-score regression represent genetic associations with no information on causal direction). This may indicate that gene expression changes contributing to chronic pain development as found in these S-PrediXcan analyses do not directly contribute to psychiatric conditions such as MDD. This would be in alignment with previous studies suggesting chronic pain can have a downstream causal effect on major depression development but that major depression does not have a causal effect on chronic pain (10). It may also be the case that tissues not examined in this study are associated with MCP-GREX and would show association with psychiatric disorder or other expected phecodes in a PheWAS setting, although it is difficult to justify why these non-brain tissues would show this result when brain tissues do not– we chose to examine brain and whole blood as chronic pain involves significant changes in the brain and spinal cord (20–23), and whole blood represents a tissue of interest due to immune components and ease of testing for e.g. potential chronic pain biomarkers. Finally, phecodes generally represent a broad category of diagnoses - for example the phecode for mood disorder (296) encompasses depression associated with MDD, bipolar disorder, and schizophrenia, and this heterogeneity could affect PheWAS results.

### Changes to PheWAS findings when adjusting for mean pain score

After adjusting our PheWAS association testing for mean pain score, results were significantly different compared to main PheWAS analyses. However, these changes appear to be driven by reduction in sample size, rather than adjustment for mean pain score, as unadjusted and adjusted analyses in the same subset of individuals showed similar results. Although NRS is a widely used pain reporting measure in clinical and research settings (136) it can change unpredictable ways over time in chronic pain (137, 138), may not be accurately reflective of treatment outcome when used alone (139), and may not be the most useful measure for identifying clinically important pain (140) or clinically important changes in pain (141). Pain NRS may not represent an ideal assessment tool in non-acute pain at the population or group level, despite some studies demonstrating stability when used to assess improvement over time in indviduals (142). Perception of pain, influencing NRS, is likely to be significantly different between those with and without chronic pain (143). For example, individuals with chronic pain may experience moderate-high levels of pain, but rate this as tolerable (144), conversely depression or depressive symptoms commonly comorbid with chronic pain could lead to reporting of higher NRS (145–147).

### Drug targets in chronic pain

Chronic pain is complex and often difficult to successfully treat. Results shown here could potentially assist in prioritizing drug targets for development of chronic pain treatments– genes where MCP-GREX is associated with upregulation may present better targets in genomic medicine (downregulation of a gene can be easier to induce than upregulation), and genes where significant MCP-GREX is shown in a small range or singular tissue may present a better target for potential animal modelling of chronic pain, compared to genes where MCP-GREX is shown to be widespread. DrugBank lookups here provide initial suggestions for potential drug repurposing in chronic pain, and in fact, several drugs highlighted here are already being explored experimentally in chronic pain treatment – one class being monoclonal antibodies for use in treating migraine (148–150). Another drug type with applications in chronic pain treatment is drugs that increase inhibitory glycinergic neurotransmission in the spinal cord (151, 152). Glycine interacts with SHMT1 (153), a gene where significant MCP-GREX was found in the hypothalamus.

## Conclusions

We carried out the largest TI study of a chronic pain trait to date, making important progress in translating GWAS findings to insights into mechanisms of chronic pain development. Genes highlighted by these analyses represent potential contributors to chronic pain development. In addition to this indication of causal direction, specific brain tissues and direction of effect of significant MCP-GREX are also given, adding to understanding of mechanisms of chronic pain development and beginning to bridge the gap between genotype (GWAS output) and phenotype (multisite chronic pain). Specific pathways of interest and potential mechanistic overlap with other medical conditions are indicated, and several genes showing significant MCP-GREX are also potential drug targets. Results of our PheWAS adjusted for mean pain score indicate that associations tend not to be driven solely by pain perception as measured by mean pain NRS. PheWAS results indicate potential shared causal pathways, particularly in metabolism and immune dysfunction, between chronic pain and conditions such as metabolic syndrome, anemias, and cardiac dysrhythmia.

## Data Availability

All data produced in the present study are available upon reasonable request to the authors. GWAS summary statistics for multisite chronic pain are available online at http://dx.doi.org/10.5525/gla.researchdata.822

## Acknowledgements

KJAJ is supported by NIMH (R01MH118278; R01MH124839). JSJ and LMH are supported by the Klarman Family Foundation. LMH is supported by NIMH (R01MH118278; R01MH124839) and the US National Institute of Environmental Health Sciences (R01ES033630).

