## Supplemental Table 1 for "Chronic pain gene expression changes in the brain and relationships with clinical traits"

PheWas results: mean painscore-adjusted. P (Raw) = unadjusted p value, P (FDR) = FDR-adjusted p value, Ncase = N cases for phecode, Nctrl = N controls for phecode.

| Gene | Phecode Description | Tissue | Zscore | P (FDR) | P (Raw) | Ncase | Nctrl |
| --- | --- | --- | --- | --- | --- | --- | --- |
| UFL1 | Pain and other symptoms associated with female genital organs | Brain Cerebellum | -4.44 | 0.0425 | 9.02E-06 | 14 | 7715 |
| SLC38A3 | Joint/ ligament sprain | Brain Caudate basal ganglia | 5.83 | 0.00001 | 5.69E-09 | 20 | 7810 |
| ERICH2 | Disc disorders | Brain Amygdala | -5.05 | 0.0015 | 4.38E-07 | 738 | 17112 |
| Novel Transcript | Spondylosis with myeolopathy | Brain Anterior cingulate cortex BA24 | 4.55 | 0.0315 | 5.34E-06 | 140 | 17813 |
| Novel Transcript | Spondylosis with myeolopathy | Brain Cortex | 4.62 | 0.0315 | 3.87E-06 | 140 | 17813 |
