## Supplemental Table 2 for "Chronic pain gene expression changes in the brain and relationships with clinical traits"

PheWas results: matched sample for mean painscore-adjusted, without adjustment for mean painscore. P (Raw) = unadjusted p value, P (FDR) = FDR-adjusted p value, Ncase = N cases for phecode, Nctrl = N controls for phecode.

| Gene | Phecode Description | Tissue | Zscore | P (FDR) | P (Raw) | Ncase | Nctrl |
| --- | --- | --- | --- | --- | --- | --- | --- |
| PTK2 | Excessive or frequent menstruation | Brain_Hypothalamus | 4.81 | 0.018 | 1.49E-06 | 393 | 17300 |
| SLC38A3 | Joint/ ligament sprain | Brain_Caudate_basal_ganglia | 5.86 | 1.10E-05 | 4.66E-09 | 20 | 7815 |
| ERICH2 | disc disorders | Brain_Amygdala | -5.18 | 0.00078 | 2.21E-07 | 738 | 17123 |
| ERICH2 | displacement of intervertebral disc | Brain_Amygdala | -4.22 | 0.043 | 2.41E-05 | 426 | 17311 |
| Novel Transcript | spondylosis with myeolopathy | Brain_Anterior_cingulate_cortex_BA24 | 4.54 | 0.033 | 5.53E-06 | 140 | 17824 |
| Novel Transcript | spondylosis with myeolopathy | Brain_Cortex | 4.61 | 0.033 | 4.05E-06 | 140 | 17824 |
