## Supplementary material for "Chronic pain gene expression changes in the brain and relationships with clinical traits": Table 1

**Table 1: Within-tissue p-value Bonferroni significance thresholds.**

| **Tissue** | **p-value Threshold** |
| --- | --- |
| Amygdala | 1.79x10^-05^ |
| Anterior cingulate cortex BA24 | 1.41 x10^-05^ |
| Caudate basal ganglia | 9.99 x10^-06^ |
| Cerebellar Hemisphere | 8.69 x10^-06^ |
| Cerebellum | 7.36 x10^-06^ |
| Cortex | 9.09 x10^-06^ |
| Frontal Cortex BA9 | 1.10 x10^-05^ |
| Hippocampus | 1.36 x10^-05^ |
| Hypothalamus | 1.37 x10^-05^ |
| Nucleus accumbens basal ganglia | 1.03 x10^-05^ |
| Putamen basal ganglia | 1.13 x10^-05^ |
| Spinal cord cervical c-1 | 1.54 x10^-05^ |
| Substantia nigra | 1.95 x10^-05^ |
| Whole Blood | 6.89 x10^-06^ |
