## Supplementary material for "Chronic pain gene expression changes in the brain and relationships with clinical traits": Table 2

**Table 2: Minimum and maximum sample sizes per ancestry group included in PheWas analyses.** Minimum and maximum sample sizes refer to the MCP-GREx-phecode association testing minimum and maximum number of participants. This varies as exclusion criteria vary per phecode.

| **Ancestry Group** | **N_min_** | **N_max_** |
| --- | --- | --- |
| East Asian | 732 | 846 |
| African American | 6105 | 7514 |
| Southeast Asian | 477 | 572 |
| Hispanic American | 8373 | 10324 |
| Native American | 69 | 83 |
| European American | 8262 | 9483 |
