## Supplementary material for "Chronic pain gene expression changes in the brain and relationships with clinical traits": Table 3

**Table 3: 89 unique genes are associated with multisite chronic pain.** P-values reaching experiment-wide significance are shown in **bold**.

| **Gene symbol** | **Tissue** | **Zscore** | **Effect Size** | **P (unadj)** |
| --- | --- | --- | --- | --- |
| ECM1 | Hippocampus | 6.43 | 0.182 | **1.24x10^-10^** |
| TARS2 | Cerebellum | 6.43 | 0.157 | **1.29 x10^-10^** |
| GPX1 | Frontal Cortex BA9 | 6.25 | 0.083 | **4.03 x10^-10^** |
| GPX1 | Cerebellar hemisphere | 6.20 | 0.113 | **5.54 x10^-10^** |
| GMPPB | Anterior cingulate cortex BA24 | 6.17 | 0.051 | **6.77 x10^-10^** |
| SNRPC | Anterior cingulate cortex BA24 | -6.17 | -0.082 | **6.95 x10^-10^** |
| GMPPB | Hypothalamus | 6.17 | 0.046 | **6.96 x10^-10^** |
| GMPPB | Caudate basal ganglia | 6.15 | 0.041 | **7.77 x10^-10^** |
| GMPPB | Cerebellum | 6.14 | 0.032 | **8.38X10^-10^** |
| GMPPB | Nucleus accumbens basal ganglia | 6.12 | 0.037 | **9.08X10^-10^** |
| GMPPB | Cerebellar hemisphere | 6.07 | 0.039 | **1.30X10^-09^** |
| CELSR3 | Amygdala | 6.02 | 0.218 | **1.77X10^-09^** |
| SEMA3B | Nucleus accumbens basal ganglia | -5.97 | -0.267 | **2.32X10^-09^** |
| GMPPB | Spinal cord cervical c1 | 5.97 | 0.038 | **2.40X10^-09^** |
| GMPPB | Whole blood | 5.95 | 0.120 | **2.70X10^-09^** |
| AMT | Hypothalamus | -5.91 | -0.110 | **3.44X10^-09^** |
| GMPPB | Cortex | 5.90 | 0.032 | **3.53X10^-09^** |
| NMT1 | Putamen basal ganglia | 5.90 | 0.075 | **3.63X10^-09^** |
| NMT1 | Anterior cingulate cortex BA24 | 5.89 | 0.120 | **3.83X10^-09^** |
| VPS33B | Whole blood | -5.84 | -0.069 | **5.35X10^-09^** |
| RP11-24H2.3 | Amygdala | -5.84 | -0.059 | **5.36X10^-09^** |
| FUBP1 | Cerebellum | -5.81 | -0.227 | **6.06X10^-09^** |
| RPRD2 | Nucleus accumbens basal ganglia | 5.81 | 0.290 | **6.10X10^-09^** |
| GMPPB | Hippocampus | 5.78 | 0.032 | **7.51X10^-09^** |
| C6orf106 (ILRUN) | Hypothalamus | 5.73 | 0.163 | **1.01X10^-08^** |
| GMPPB | Substantia nigra | 5.69 | 0.031 | **1.24X10^-08^** |
| UHRF1BP1 | Spinal cord cervical c1 | 5.66 | 0.070 | **1.49X10^-08^** |
| CSK | Caudate basal ganglia | -5.58 | -0.138 | **2.35X10^-08^** |
| SNRPC | Frontal Cortex BA9 | -5.54 | -0.065 | **3.04X10^-08^** |
| AMT | Whole blood | -5.51 | -0.054 | **3.50X10^-08^** |
| GPX1 | Cortex | 5.47 | 0.063 | **4.49X10^-08^** |
| SDCCAG8 | Whole blood | 5.44 | 0.043 | **5.31X10^-08^** |
| C6orf106 (ILRUN) | Putamen basal ganglia | 5.43 | 0.068 | **5.60X10^-08^** |
| ECM1 | Nucleus accumbens basal ganglia | 5.43 | 0.101 | **5.70X10^-08^** |
| ZNF501 | Frontal Cortex BA9 | -5.41 | -0.066 | **6.26X10^-08^** |
| RBM6 | Nucleus accumbens basal ganglia | -5.40 | -0.049 | **6.68X10^-08^** |
| SNRPC | Nucleus accumbens basal ganglia | -5.37 | -0.038 | **7.95X10^-08^** |
| AMT | Putamen basal ganglia | -5.36 | -0.050 | **8.10X10^-08^** |
| C6orf106 (ILRUN) | Cortex | 5.36 | 0.078 | **8.50X10^-08^** |
| SUOX | Whole blood | 5.34 | 0.073 | **9.14X10^-08^** |
| C6orf106 (ILRUN) | Frontal Cortex BA9 | 5.33 | 0.120 | **9.78X10^-08^** |
| UHRF1BP1 | Hypothalamus | 5.30 | 0.072 | **1.14X10^-07^** |
| RP11-24H2.3 | Anterior cingulate cortex BA24 | -5.30 | -0.043 | **1.18X10^-07^** |
| MST1 | Whole blood | -5.30 | -0.125 | **1.18X10^-07^** |
| GMPPB | Frontal Cortex BA9 | 5.29 | 0.034 | **1.19X10^-07^** |
| RPS26 | Frontal Cortex BA9 | -5.28 | -0.017 | **1.30X10^-07^** |
| RNF123 | Nucleus accumbens basal ganglia | 5.27 | 0.119 | **1.36X10^-07^** |
| RPS26 | Putamen basal ganglia | -5.24 | -0.014 | **1.64X10^-07^** |
| AMT | Substantia nigra | -5.23 | -0.077 | **1.68X10^-07^** |
| GPX1 | Cerebellum | 5.23 | 0.060 | **1.69X10^-07^** |
| GPR27 | Cortex | 5.19 | 0.183 | **2.06X10^-07^** |
| C6orf106 (ILRUN) | Nucleus accumbens basal ganglia | 5.19 | 0.079 | **2.10X10^-07^** |
| SNRPC | Hippocampus | -5.19 | -0.086 | **2.13X10^-07^** |
| AMT | Anterior cingulate cortex BA24 | -5.19 | -0.075 | **2.13X10^-07^** |
| SUOX | Nucleus accumbens basal ganglia | 5.18 | 0.054 | **2.18X10^-07^** |
| UHRF1BP1 | Cerebellar hemisphere | 5.18 | 0.068 | **2.18X10^-07^** |
| MRPS21 | Frontal Cortex BA9 | -5.18 | -0.182 | **2.27X10^-07^** |
| SNRPC | Putamen basal ganglia | -5.16 | -0.054 | **2.50X10^-07^** |
| RPS26 | Cerebellum | -5.15 | -0.015 | **2.58X10^-07^** |
| RPRD2 | Whole blood | 5.13 | 0.250 | **2.87X10^-07^** |
| SNRPC | Whole blood | -5.13 | -0.486 | **2.91X10^-07^** |
| GPX1 | Caudate basal ganglia | 5.12 | 0.078 | **2.98X10^-07^** |
| UHRF1BP1 | Cortex | 5.12 | 0.044 | **3.07X10^-07^** |
| CEP170 | Whole blood | 5.10 | 0.159 | **3.34X10^-07^** |
| SUOX | Putamen basal ganglia | 5.10 | 0.079 | **3.40X10^-07^** |
| GMPPB | Amygdala | 5.09 | 0.027 | **3.55X10^-07^** |
| AMT | Nucleus accumbens basal ganglia | -5.09 | -0.049 | **3.65X10^-07^** |
| SDCCAG8 | Caudate basal ganglia | 5.08 | 0.114 | **3.70X10^-07^** |
| P4HTM | Cerebellum | -5.08 | -0.072 | **3.83X10^-07^** |
| RBM6 | Caudate basal ganglia | -5.06 | -0.050 | **4.14X10^-07^** |
| INTS1 | Spinal cord cervical c1 | -5.06 | -0.025 | **4.19X10^-07^** |
| RBM6 | Cortex | -5.06 | -0.031 | **4.27X10^-07^** |
| RP11-160H22.5 | Whole blood | -5.06 | -0.059 | **4.28X10^-07^** |
| UHRF1BP1 | Caudate basal ganglia | 5.04 | 0.062 | **4.59X10^-07^** |
| UHRF1BP1 | Whole blood | 5.04 | 0.029 | **4.71X10^-07^** |
| SUOX | Cerebellum | 5.03 | 0.027 | **4.88X10^-07^** |
| SUOX | Cerebellar hemisphere | 5.03 | 0.044 | **4.90X10^-07^** |
| UHRF1BP1 | Frontal Cortex BA9 | 5.02 | 0.157 | **5.17X10^-07^** |
| SP4 | Nucleus accumbens basal ganglia | 5.00 | 0.243 | **5.61X10^-07^** |
| MON1B | Whole blood | 5.00 | 0.105 | **5.63X10^-07^** |
| SUOX | Caudate basal ganglia | 5.00 | 0.075 | **5.82X10^-07^** |
| SDCCAG8 | Putamen basal ganglia | 4.99 | 0.087 | **5.89X10^-07^** |
| RPRD2 | Substantia nigra | 4.99 | 0.102 | **5.91X10^-07^** |
| ZNF197 | Whole blood | -4.99 | -0.037 | **5.98X10^-07^** |
| GPR27 | Frontal Cortex BA9 | 4.98 | 0.137 | **6.21X10^-07^** |
| UHRF1BP1 | Cerebellum | 4.98 | 0.080 | **6.28X10^-07^** |
| CTBP2 | Cerebellum | -4.98 | -0.080 | **6.40X10^-07^** |
| GPX1 | Nucleus accumbens basal ganglia | 4.96 | 0.055 | **7.03X10^-07^** |
| PTK2 | Nucleus accumbens basal ganglia | 4.95 | 0.079 | **7.56X10^-07^** |
| SLC25A13 | Whole blood | 4.94 | 0.070 | **7.66X10^-07^** |
| RPS26 | Caudate basal ganglia | -4.94 | -0.016 | **7.80X10^-07^** |
| SEMA3F | Anterior cingulate cortex BA24 | -4.94 | -0.205 | **7.80X10^-07^** |
| RPS26 | Nucleus accumbens basal ganglia | -4.94 | -0.014 | **7.89X10^-07^** |
| RNF123 | Cerebellum | 4.94 | 0.042 | **7.94X10^-07^** |
| SUOX | Cortex | 4.94 | 0.071 | **7.94X10^-07^** |
| SUOX | Amygdala | 4.94 | 0.081 | 7.99X10^-07^ |
| SUOX | Hypothalamus | 4.93 | 0.073 | 8.10X10^-07^ |
| RBM6 | Cerebellar hemisphere | -4.92 | -0.044 | 8.50X10^-07^ |
| RPS26 | Whole blood | -4.92 | -0.013 | 8.54X10^-07^ |
| SP4 | Cerebellum | 4.92 | 0.265 | 8.85X10^-07^ |
| MRPS21 | Cerebellum | -4.92 | -0.048 | 8.86X10^-07^ |
| NUDT18 | Putamen basal ganglia | -4.91 | -0.062 | 8.95X10^-07^ |
| MRPS21 | Hypothalamus | -4.91 | -0.181 | 8.99X10^-07^ |
| GMPPB | Putamen basal ganglia | 4.91 | 0.032 | 9.01X10^-07^ |
| SUOX | Spinal cord cervical c1 | 4.91 | 0.062 | 9.02X10^-07^ |
| RPS26 | Cortex | -4.90 | -0.016 | 9.78X10^-07^ |
| TARS2 | Anterior cingulate cortex BA24 | 4.89 | 0.141 | 9.99X10^-07^ |
| RNF123 | Cortex | 4.88 | 0.093 | 1.05X10^-06^ |
| UHRF1BP1 | Putamen basal ganglia | 4.88 | 0.032 | 1.07X10^-06^ |
| RBM6 | Frontal Cortex BA9 | -4.88 | -0.050 | 1.08X10^-06^ |
| AMT | Cortex | -4.87 | -0.033 | 1.09X10^-06^ |
| RPS26 | Hypothalamus | -4.87 | -0.016 | 1.11X10^-06^ |
| GPX1 | Putamen basal ganglia | 4.87 | 0.118 | 1.14X10^-06^ |
| NUDT18 | Whole blood | -4.86 | -0.036 | 1.14X10^-06^ |
| RBM6 | Whole blood | -4.86 | -0.025 | 1.15X10^-06^ |
| RBM6 | Putamen basal ganglia | -4.86 | -0.039 | 1.15X10^-06^ |
| RPS26 | Cerebellar hemisphere | -4.86 | -0.017 | 1.15X10^-06^ |
| PRKAR2A | Substantia nigra | 4.86 | 0.099 | 1.16X10^-06^ |
| ECM1 | Cerebellar hemisphere | 4.85 | 0.033 | 1.25X10^-06^ |
| RPS26 | Anterior cingulate cortex BA24 | -4.84 | -0.014 | 1.27X10^-06^ |
| MRPS21 | Caudate basal ganglia | -4.84 | -0.090 | 1.29X10^-06^ |
| UFL1 | Cerebellum | 4.84 | 0.068 | 1.31X10^-06^ |
| ZNF501 | Caudate basal ganglia | -4.82 | -0.059 | 1.41X10^-06^ |
| SCAMP2 | Cerebellum | 4.81 | 0.093 | 1.48X10^-06^ |
| MRPS21 | Nucleus accumbens basal ganglia | -4.81 | -2.206 | 1.50X10^-06^ |
| NMT1 | Caudate basal ganglia | 4.81 | 0.080 | 1.54X10^-06^ |
| TSKU | Cerebellar hemisphere | 4.80 | 0.038 | 1.55X10^-06^ |
| UBA7 | Caudate basal ganglia | -4.80 | -0.240 | 1.56X10^-06^ |
| LANCL1 | Cortex | 4.80 | 0.102 | 1.59X10^-06^ |
| GRK4 | Anterior cingulate cortex BA24 | 4.80 | 0.060 | 1.62X10^-06^ |
| ZNF501 | Cerebellum | -4.79 | -0.046 | 1.71X10^-06^ |
| UHRF1BP1 | Nucleus accumbens basal ganglia | 4.78 | 0.149 | 1.73X10^-06^ |
| SNRPC | Caudate basal ganglia | -4.78 | -0.059 | 1.77X10^-06^ |
| C15orf57 | Cortex | -4.78 | -0.035 | 1.79X10^-06^ |
| UHRF1BP1 | Anterior cingulate cortex BA24 | 4.78 | 0.058 | 1.79X10^-06^ |
| RBM6 | Anterior cingulate cortex BA24 | -4.77 | -0.035 | 1.80X10^-06^ |
| MST1R | Caudate basal ganglia | 4.77 | 0.068 | 1.82X10^-06^ |
| KLHDC8B | Cerebellum | 4.77 | 0.113 | 1.86X10^-06^ |
| TSPYL4 | Cerebellum | 4.76 | 0.099 | 1.93X10^-06^ |
| C15orf57 | Nucleus accumbens basal ganglia | -4.76 | -0.040 | 1.93X10^-06^ |
| ZNF35 | Whole blood | -4.76 | -0.127 | 1.93X10^-06^ |
| RBM6 | Spinal cord cervical c1 | -4.75 | -0.043 | 2.07X10^-06^ |
| LIN28B-AS1 | Putamen basal ganglia | 4.73 | 0.120 | 2.24X10^-06^ |
| AMT | Caudate basal ganglia | -4.73 | -0.031 | 2.29X10^-06^ |
| MAU2 | Cerebellum | -4.72 | -0.124 | 2.31X10^-06^ |
| TSKU | Cerebellum | 4.71 | 0.044 | 2.48X10^-06^ |
| RPS26 | Amygdala | -4.71 | -0.015 | 2.53X10^-06^ |
| SNRPC | Amygdala | -4.69 | -0.045 | 2.68X10^-06^ |
| ACADL | Frontal Cortex BA9 | -4.69 | -0.208 | 2.71X10^-06^ |
| PACSIN3 | Cortex | -4.69 | -0.095 | 2.72X10^-06^ |
| C6orf106 (ILRUN) | Amygdala | 4.68 | 0.089 | 2.80X10^-06^ |
| MPI | Putamen basal ganglia | 4.68 | 0.058 | 2.81X10^-06^ |
| PTK2 | Caudate basal ganglia | 4.68 | 0.097 | 2.85X10^-06^ |
| NUP43 | Cerebellum | -4.68 | -0.033 | 2.93X10^-06^ |
| KNDC1 | Cerebellum | 4.68 | 0.035 | 2.94X10^-06^ |
| NUP43 | Cerebellar hemisphere | -4.67 | -0.037 | 3.04X10^-06^ |
| RBM6 | Hippocampus | -4.67 | -0.071 | 3.08X10^-06^ |
| SNRPC | Cortex | -4.66 | -0.036 | 3.15X10^-06^ |
| GINM1 | Whole blood | 4.66 | 0.054 | 3.17X10^-06^ |
| FASTKD5 | Cortex | 4.66 | 0.108 | 3.20X10^-06^ |
| UBOX5 | Nucleus accumbens basal ganglia | -4.65 | -0.080 | 3.27X10^-06^ |
| AMT | Hippocampus | -4.65 | -0.049 | 3.36X10^-06^ |
| HEXIM1 | Frontal Cortex BA9 | -4.65 | -0.129 | 3.37X10^-06^ |
| KCNH2 | Cerebellar hemisphere | -4.64 | -0.057 | 3.46X10^-06^ |
| NELFA | Cerebellum | 4.64 | 0.097 | 3.47X10^-06^ |
| P4HTM | Cerebellar hemisphere | -4.64 | -0.065 | 3.50X10^-06^ |
| ERICH2 | Amygdala | -4.63 | -0.072 | 3.74X10^-06^ |
| RNF123 | Cerebellar hemisphere | 4.60 | 0.055 | 4.30X10^-06^ |
| LATS1 | Cerebellum | -4.59 | -0.067 | 4.51X10^-06^ |
| RNF123 | Amygdala | 4.58 | 0.106 | 4.65X10^-06^ |
| DCAKD | Frontal Cortex BA9 | -4.58 | -0.052 | 4.68X10^-06^ |
| NUDT18 | Amygdala | -4.58 | -0.186 | 4.69X10^-06^ |
| DCAKD | Whole blood | -4.58 | -0.044 | 4.75X10^-06^ |
| RBM6 | Cerebellum | -4.57 | -0.022 | 4.80X10^-06^ |
| C6orf106 (ILRUN) | Cerebellar hemisphere | 4.57 | 0.115 | 4.85X10^-06^ |
| RNF123 | Anterior cingulate cortex BA24 | 4.57 | 0.102 | 4.87X10^-06^ |
| AC007405.6 | Caudate basal ganglia | -4.57 | -0.080 | 4.93X10^-06^ |
| NUDT18 | Nucleus accumbens basal ganglia | -4.57 | -0.033 | 4.98X10^-06^ |
| PPP6C | Anterior cingulate cortex BA24 | 4.56 | 0.109 | 5.04X10^-06^ |
| LLGL1 | Anterior cingulate cortex BA24 | -4.56 | -0.222 | 5.07X10^-06^ |
| NUP43 | Whole blood | 4.56 | 0.043 | 5.19X10^-06^ |
| C15orf57 | Cerebellum | -4.55 | -0.028 | 5.42X10^-06^ |
| ZNF23 | Hippocampus | 4.54 | 0.049 | 5.51X10^-06^ |
| RPS26 | Substantia nigra | -4.54 | -0.017 | 5.54X10^-06^ |
| PPP6C | Cortex | 4.54 | 0.289 | 5.58X10^-06^ |
| SLC38A3 | Frontal Cortex BA9 | -4.54 | -0.065 | 5.62X10^-06^ |
| ZNF502 | Hippocampus | -4.54 | -0.081 | 5.63X10^-06^ |
| DNAH11 | Frontal Cortex BA9 | 4.54 | 0.093 | 5.68X10^-06^ |
| ZNF502 | Nucleus accumbens basal ganglia | -4.54 | -0.140 | 5.71X10^-06^ |
| SCAMP2 | Whole blood | 4.54 | 0.088 | 5.75X10^-06^ |
| RAD51 | Caudate basal ganglia | -4.53 | -0.070 | 5.81X10^-06^ |
| ZNF502 | Cortex | -4.52 | -0.029 | 6.05X10^-06^ |
| DCAKD | Cerebellum | -4.52 | -0.027 | 6.25X10^-06^ |
| URM1 | Whole blood | -4.52 | -0.148 | 6.32X10^-06^ |
| LATS1 | Caudate basal ganglia | -4.51 | -0.151 | 6.36X10^-06^ |
| BAK1 | Cerebellum | 4.51 | 0.071 | 6.40X10^-06^ |
| NUDT18 | Caudate basal ganglia | -4.50 | -0.059 | 6.71X10^-06^ |
| MPI | Anterior cingulate cortex BA24 | 4.50 | 0.037 | 6.76X10^-06^ |
| FAM180B | Hypothalamus | -4.50 | -0.052 | 6.77X10^-06^ |
| IL23A | Hypothalamus | 4.50 | 0.059 | 6.85X10^-06^ |
| ZNF502 | Whole blood | -4.50 | -0.034 | 6.86X10^-06^ |
| DNMT3B | Cerebellum | 4.50 | 0.047 | 6.93X10^-06^ |
| LANCL1 | Cerebellar hemisphere | 4.49 | 0.113 | 6.97X10^-06^ |
| MPI | Cerebellum | 4.49 | 0.079 | 6.97X10^-06^ |
| SCAI | Cortex | 4.49 | 0.133 | 7.06X10^-06^ |
| SLC25A13 | Cerebellar hemisphere | 4.48 | 0.058 | 7.29X10^-06^ |
| CDK14 | Cortex | 4.48 | 0.164 | 7.36X10^-06^ |
| ACSF3 | Cortex | -4.47 | -0.023 | 7.73X10^-06^ |
| KIF3B | Amygdala | 4.47 | 0.064 | 7.81X10^-06^ |
| RP11-147L13.8 | Frontal Cortex BA9 | 4.47 | 0.058 | 7.96X10^-06^ |
| RP11-147L13.11 | Spinal cord cervical c1 | -4.46 | -0.146 | 8.16X10^-06^ |
| RNF123 | Hypothalamus | 4.46 | 0.106 | 8.30X10^-06^ |
| MST1R | Nucleus accumbens basal ganglia | 4.46 | 0.041 | 8.34X10^-06^ |
| LINC01671 | Nucleus accumbens basal ganglia | -4.45 | -0.090 | 8.42X10^-06^ |
| CYB561D2 | Cortex | -4.45 | -0.172 | 8.44X10^-06^ |
| S100A1 | Cortex | 4.45 | 0.163 | 8.58X10^-06^ |
| RBM6 | Hypothalamus | -4.44 | -0.043 | 8.96X10^-06^ |
| RBM6 | Substantia nigra | -4.41 | -0.035 | 1.01X10^-05^ |
| DNAH11 | Putamen basal ganglia | 4.41 | 0.045 | 1.03X10^-05^ |
| C15orf57 | Hypothalamus | -4.40 | -0.032 | 1.06X10^-05^ |
| ZNF502 | Frontal Cortex BA9 | -4.40 | -0.041 | 1.10X10^-05^ |
| COX11 | Anterior cingulate cortex BA24 | -4.39 | -0.049 | 1.11X10^-05^ |
| NMT1 | Hippocampus | 4.39 | 0.085 | 1.13X10^-05^ |
| BAK1 | Hippocampus | 4.39 | 0.070 | 1.15X10^-05^ |
| SHMT1 | Hypothalamus | 4.38 | 0.044 | 1.18X10^-05^ |
| COX11 | Amygdala | -4.37 | -0.059 | 1.22X10^-05^ |
| COX11 | Hippocampus | -4.37 | -0.069 | 1.27X10^-05^ |
| RP11-147L13.11 | Anterior cingulate cortex BA24 | -4.36 | -0.102 | 1.30X10^-05^ |
| GINM1 | Substantia nigra | 4.36 | 0.076 | 1.31X10^-05^ |
