## Supplementary material for "Chronic pain gene expression changes in the brain and relationships with clinical traits": Table 4

**Table 4: DrugBank lookup results for 89 significant MCP-GREX genes.**

PD = Parkinson disease, AD = Alzheimer disease, CFS = chronic fatigue syndrome, CVD = cardiovascular disease, OCD = obsessive compulsive disorder. Genes in **bold** were also found to be significant in PheWas analyses.

| **Symbol** | **DrugBank Accession ID** | **Drug Name & Description** |
| --- | --- | --- |
| AMT | DB00116 | tetrahydrofolic acid, nutritional supplement |
|  | DB00157 | NADH, nutritional supplement (some evidence for PD, CFS, AD, CVD benefit) |
|  | DB04789 | 5-methyltetrahydrofolic acid, nutritional supplement |
| CSK | DB01254 | dasatinib, tyrosine kinase inhibitor, cancer treatment (leukemia) |
|  | DB02010 | staurosporine, protein kinase C inhibitor |
|  | DB05075 | TG-100801, topically applied kinase inhibitor (macular degeneration) |
|  | DB12010 | fostamatinib, spleen tyrosine kinase inhibitor (thrombocytopenia) |
| FUBP1 | DB05786 | irofulven, novel anti-cancer compound |
| GPX1 | DB00143 | glutathione, nutritional supplement |
| IL23A | DB05459 | briakinumab, anti-IL-12 monoclonal antibody for T-cell-driven autoimmune disease treatment |
|  | DB11834 | guselkumab, monclonal antibody for plaque psoriasis |
| KCNH2 | DB00176 | fluvoxamine, SSRI for OCD |
|  | DB00199 | erythromycin, macrolide antibiotic |
|  | DB00204 | dofetilide, class 3 antiarrhythmic |
|  | DB00276 | amsacrine, cytotoxin for leukemia treatment |
|  | DB00280 | disopyramide, class 1A antiarrhythmic |
|  | DB00308 | ibutilide, class 3 antiarrythmic |
|  | DB00342 | terfenadine, antihistamine |
|  | DB00346 | alfuzosin, alpha-1 adrenergic antagonist |
|  | DB00455 | loratidine, 2nd generation antihistamine |
|  | DB00457 | prazosin, alpha-blocker for hypertension |
|  | DB00458 | imipramine, tricyclic antidepressant |
|  | DB00472 | fluoxetine, SSRI |
|  | DB00477 | chlorpromazine, phenothiazine antipsychotic |
|  | DB00489 | sotalol, methane sulfoanilide beta adrenergic antagonist for arrhythmia |
|  | DB00537 | ciprofloxacin, second generation fluoroquinolone |
|  | DB00590 | doxazosin, alpha-1 adrenergic receptor for hypertension |
|  | DB00604 | cisapride, GERD-associated heartburn medication |
|  | DB00637 | astemizole, second generation antihistamine |
|  | DB00661 | verapamil, non-dihydropyridine calcium channel blocker for angina, arrhythmia, hypertension |
|  | DB00675 | tamoxifen, selective estrogen receptor modulator used in certain breast cancers |
|  | DB00679 | thioridazine, phenothiazine antipsychotic for GAD and schizophrenia |
|  | DB00908 | quinidine, arrhythmia treatment |
|  | DB01026 | ketoconazole, broad spectrum antifungal |
|  | DB01035 | procainamide, arrhythmia treatment |
|  | DB01074 | perhexiline, coronary vasodilator |
|  | DB01100 | pimozide, antipsychotic used in Tourette's |
|  | DB01110 | miconazole, azole antifungal |
|  | DB01118 | amiodarone, class 3 antiarrhythmic |
|  | DB01136 | carvedilol, non-selective beta-adrenergic antagonist |
|  | DB01142 | doxepin, psychotropic agent with antidepressant and anxiolytic properties |
|  | DB01149 | nefazodone, antidepressant |
|  | DB01162 | terazosin, alpha-1 adrenergic antagonist |
|  | DB01182 | propafenone, class 1c antiarrhythmic |
|  | DB01195 | flecainide, class 1c antiarrhythmic |
|  | DB01211 | clarithromycin, macrolide antibiotic |
|  | DB01218 | halofantrine, antimalarial |
|  | DB01244 | bepridil, calcium channel blocker |
|  | DB04855 | dronedarone, antiarrhythmic |
|  | DB04957 | azimilide, investigational class 3 antiarrhythmic |
|  | DB06144 | sertindole, atypical antipsychotic |
|  | DB06217 | vernakalant, antiarrhythmic |
|  | DB06457 | tecastemizole, investigational small molecule |
|  | DB11090 | potassium nitrate, small wound cauterization |
|  | DB11186 | pentoxyverine, cough suppressant |
|  | DB11386 | chlorobutanol, alcohol-based perservative |
|  | DB11633 | Isavuconazole, triazole antifungal |
|  | DB11642 | pitolisant, antagonist of histamine H3 receptor, narcolepsy treatment |
| LATS1 | DB12010 | fostamatinib, spleen tyrosine kinase inhibitor (thrombocytopenia) |
| MST1R | DB12010 | fostamatinib, spleen tyrosine kinase inhibitor (thrombocytopenia) |
| NMT1 | DB03062 | investigational small molecule |
| P4HTM | DB00126 | vitamin C, nutritional supplement |
| PRKAR2A | DB05798 | GEM-231, monoclonal antibody |
| PTK2 | DB06423 | endostatin, investigational small molecule |
|  | DB07248 | investigational small molecule |
|  | DB07460 | investigational small molecule |
|  | DB12010 | fostamatinib, spleen tyrosine kinase inhibitor (thrombocytopenia) |
| **RAD51** | DB04395 | phosphoaminophosphonic acid-adenylate ester, investigational small molecule |
|  | DB12742 | amuvatinib, cancer treatment undergoing clinical trial |
| S100A1 | DB00768 | olopatadine, histamine H1 antagonist |
| SHMT1 | DB00114 | pyridoxal phosphate (B6), nutritional supplement |
|  | DB00116 | tetrahydrofolic acid, nutritional supplement |
|  | DB00145 | glycine, total parenteral nutrition component |
|  | DB01055 | mimosine, antineoplastic |
|  | DB02067 | triglu-5-formyl-tetrahydrofolate, investigational small molecule |
|  | DB02800 | 5-hydroxymethyl-5,6-dihydrofolic acid, investigational small molecule |
|  | DB02824 | N-pyridoxyl-glycine-5-monophosphate, investigational small molecule |
| SLC25A13 | DB00128 | aspartic acid, total parenteral nutrition component |
| **SLC38A3** | DB00117 | histidine, total parenteral nutrition component |
|  | DB00174 | asparagine, non-essential amino acid |
| SUOX | DB03983 | investigational small molecule |
| TARS2 | DB00156 | threonine, total parenteral nutrition component |
