## Supplementary material for "Chronic pain gene expression changes in the brain and relationships with clinical traits": Table 5

**Table 5: Associations between mean pain score and MCP-GREx.**

| **Gene** | **Tissue** | **Zscore** | **P (FDR)** | **P (Raw)** |
| --- | --- | --- | --- | --- |
| SDCCAG8 | Brain_Cerebellar_Hemisphere | -2.25 | 0.049 | 0.025 |
|  | Brain_Putamen_basal_ganglia | -2.65 | 0.043 | 0.008 |
|  | Whole_Blood | -2.45 | 0.043 | 0.014 |
| UHRF1BP1 | Brain_Amygdala | -3.58 | 0.002 | 0.000 |
|  | Brain_Anterior_cingulate_cortex_BA24 | -2.78 | 0.011 | 0.005 |
|  | Brain_Caudate_basal_ganglia | -2.72 | 0.011 | 0.006 |
|  | Brain_Cerebellar_Hemisphere | -2.68 | 0.011 | 0.007 |
|  | Brain_Cerebellum | -2.63 | 0.011 | 0.009 |
|  | Brain_Cortex | -3.56 | 0.002 | 0.000 |
|  | Brain_Frontal_Cortex_BA9 | -2.67 | 0.011 | 0.008 |
|  | Brain_Hypothalamus | -2.71 | 0.011 | 0.007 |
|  | Brain_Nucleus_accumbens_basal_ganglia | -2.90 | 0.011 | 0.004 |
|  | Brain_Putamen_basal_ganglia | -2.35 | 0.020 | 0.019 |
|  | Brain_Spinal_cord_cervical_c-1 | -3.03 | 0.011 | 0.002 |
|  | Whole_Blood | -2.52 | 0.014 | 0.012 |
| DNMT3B | Brain_Anterior_cingulate_cortex_BA24 | -2.91 | 0.011 | 0.004 |
| ACADL | Brain_Frontal_Cortex_BA9 | 2.42 | 0.031 | 0.015 |
| SNRPC | Brain_Amygdala | 2.95 | 0.017 | 0.003 |
|  | Brain_Caudate_basal_ganglia | 2.13 | 0.046 | 0.033 |
|  | Brain_Cerebellum | 2.69 | 0.017 | 0.007 |
|  | Brain_Cortex | 2.67 | 0.017 | 0.008 |
|  | Brain_Hippocampus | 2.69 | 0.017 | 0.007 |
|  | Brain_Nucleus_accumbens_basal_ganglia | 2.50 | 0.021 | 0.012 |
|  | Brain_Putamen_basal_ganglia | 2.48 | 0.021 | 0.013 |
|  | Whole_Blood | 3.02 | 0.017 | 0.003 |
| TARS2 | Brain_Anterior_cingulate_cortex_BA24 | 3.26 | 0.006 | 0.001 |
|  | Brain_Cerebellar_Hemisphere | 2.39 | 0.028 | 0.017 |
|  | Brain_Cerebellum | 2.98 | 0.007 | 0.003 |
| CEP170 | Whole_Blood | -2.52 | 0.023 | 0.012 |
| HEXIM1 | Brain_Cortex | 2.60 | 0.028 | 0.009 |
| ILRUN | Brain_Hippocampus | -3.07 | 0.024 | 0.002 |
| MRPS21 | Brain_Caudate_basal_ganglia | 2.48 | 0.025 | 0.013 |
|  | Brain_Cerebellum | -3.15 | 0.015 | 0.002 |
|  | Brain_Cortex | -2.45 | 0.025 | 0.014 |
|  | Brain_Frontal_Cortex_BA9 | -2.53 | 0.025 | 0.011 |
|  | Brain_Hypothalamus | -2.37 | 0.027 | 0.018 |
|  | Brain_Nucleus_accumbens_basal_ganglia | 2.71 | 0.025 | 0.007 |
