## Supplementary material for "Chronic pain gene expression changes in the brain and relationships with clinical traits": Table 6

**Table 6: Significant GREX-phecode associations.**

FDR = false-discovery rate correction (carried out within-gene). Gene = gene symbol, Zscore = PheWas Z score value, tissue = GREX tissue, P (FDR) = GREX-phecode PheWas p value (FDR-corrected), P (Raw) = uncorrected p value.

| **Gene** | **Phecode Description** | **Tissue** | **Full Analysis** | | | **Correcting for Pain Scores** | | |
| --- | --- | --- | --- | --- | --- | --- | --- | --- |
|  |  |  | **Z-score** | **P (FDR)** | **P (Raw)** | **Z-score** | **P (FDR)** | **P (Raw)** |
| DCAKD | Cardiac dysrhythmias | Caudate Basal Ganglia | -4.98 | 0.0084 | 6.18x10-7 | -5.52 | 0.0046 | 3.47x10^-8^ |
| ECM1 | Dysmetabolic syndrome x | Cerebellar Hemisphere | 4.58 | 0.0063 | 6.39x10-7 |  |  |  |
|  |  | Cerebellum | -4.99 | 0.0228 | 4.61x10-6 |  |  |  |
|  |  | Nucleus accumbens basal ganglia | 5.33 | 0.026 | 7.89x10-6 |  |  |  |
| ERICH2 | Disc disorders/ dorsopathies | Amygdala | 4.7 | 0.0312 | 8.4x10-6 | -4.42 | 0.033 | 9.83x10^-06^ |
| ILRUN (a.k.a. C6orf106) | Primary thrombocytopenia | Amygdala | -4.47 | 0.0176 | 1.98x10-6 |  |  |  |
|  |  | Hypothalamus | 4.81 | 0.0176 | 2.59x10-6 |  |  |  |
|  |  | Nucleus accumbens basal ganglia | 4.46 | 0.024 | 5.3x10-6 |  |  |  |
|  |  | Cortex | -4.45 | 0.0415 | 1.22x10-5 |  |  |  |
| MON1B | Anemias | Spinal cord cervical c-1 | -4.58 | 0.014 | 1.14x10-6 |  |  |  |
|  |  | Amygdala | 4.21 | 0.0293 | 4.74x10-6 |  |  |  |
| PACSIN3 | Bullous dermatoses | Nucleus accumbens basal ganglia | 4.87 | 0.0076 | 1.54x10-6 |  |  |  |
| RAD51 | Disturbances of sulphur-bearing amino-acid metabolism | Substantia Nigra | 4.76 | 0.0307 | 1.24x10-5 |  |  |  |
| SCAI | Inflammatory and toxic neuropathy | Cortex | 4.55 | 0.0412 | 8.34x10-6 |  |  |  |
| SLC38A3 | Joint/ ligament sprain | Caudate basal ganglia | 4.37 | 0.0002 | 9.62x10-8 | 6.00 | 4.34x10^-06^ | 1.92x10^-09^ |
|  | Neurological disorders | Caudate basal ganglia | 4.37 | 0.0309 | 2.5x10-5 |  |  |  |
| ZNF197 | Hand/ finger injuries and lacerations | Substantia nigra |  |  |  | 4.45 | 0.048 | 8.45x10^-06^ |
| ENSG00000278730 (Novel transcript, lncRNA, a.k.a. RP11-147L13.11) | Spondylosis with myeolopathy | Anterior cingulate cortex BA24 |  |  |  | 4.93 | 0.0046 | 8.21x10^-07^ |
|  |  | Cerebellum |  |  |  | 4.59 | 0.017 | 4.41x10^-06^ |
|  |  | Cortex |  |  |  | 5.05 | 0.0046 | 4.46x10^-07^ |
|  |  | Hypothalamus |  |  |  | 4.37 | 0.035 | 1.25x10^-05^ |
